# Clinical Acceptability of Automatically Generated Lymph Node Levels and Structures of Deglutition and Mastication for Head and Neck Cancer Patient Radiation Treatment Planning

**DOI:** 10.1101/2023.08.07.23293787

**Authors:** Sean Maroongroge, Abdallah Sherif Radwan Mohamed, Callistus Nguyen, Jean Guma De la Vega, Steven J. Frank, Adam S. Garden, Brandon Gunn, Anna Lee, Lauren L. Mayo, Amy C. Moreno, William H. Morrison, Jack Phan, Michael T. Spiotto, Laurence E. Court, Clifton D. Fuller, David I. Rosenthal, Tucker J. Netherton

## Abstract

**Purpose/Objective(s):** Here we investigate an approach to develop and clinically validate auto-contouring models for lymph node levels and structures of deglutition and mastication in the head and neck. An objective of this work is to provide high quality resources to the scientific community to promote advancement of treatment planning, clinical trial management, and toxicity studies for the head and neck.

**Materials/Methods:** CTs of 145 patients who were irradiated for a head and neck primary malignancy at MD Anderson Cancer Center were retrospectively curated. Data were contoured by radiation oncologists and a resident physician and divided into two separate cohorts. One cohort was used to analyze lymph node levels (IA, IB, II, III, IV, V, RP) and the other used to analyze 17 swallowing and chewing structures. Forty-seven patients were in the lymph node level cohort (training/testing = 32/15). All these patients received definitive radiotherapy without a nodal dissection to minimize anatomic perturbation of the lymph node levels. The remaining 98 patients formed the swallowing/chewing structures cohort (training/testing =78/20). Separate nnUnet models were trained and validated using the separate cohorts. For the lymph node levels, two double blinded studies were used to score preference and clinical acceptability (using a 5-point Likert scale) of AI vs human contours. For the swallowing and chewing structures, clinical acceptability was scored. Quantitative analyses of the test sets were performed for AI vs human contours for all structures using the Dice Similarity Coefficient (DSC) and the 95^208^ percentile Hausdorff distance (HD95th).

**Results:** Across all lymph node levels (IA, IB, II, III, IV, V, RP), median DSC ranged from 0.77 to 0.89 for AI vs manual contours in the testing cohort. Across all lymph node levels, the AI contour was superior to or equally preferred to the manual contours at rates ranging from 75% to 91% in the first blinded study. In the second blinded study, physician preference for the manual vs AI contour was statistically different for only the RP contours (p < 0.01). Thus, there was not a significant difference in clinical acceptability for nodal levels I-V for manual versus AI contours. Across all physician-generated contours, 82% were rated as usable with stylistic to no edits, and across all AI-generated contours, 92% were rated as usable with stylistic to no edits. For the swallowing structures median DSC ranged from 0.86 to 0.96 and was greater than 0.90 for 11/17 structures types. Of the 340 contours in the test set, only 4% required minor edits.

**Conclusions:** An approach to generate clinically acceptable automated contours for lymph node levels and swallowing and chewing structures in the head and neck was demonstrated. For nodal levels I-V, there was no significant difference in clinical acceptability in manual vs AI contours. Of the two testing cohorts for lymph nodes and swallowing and chewing structures, only 8% and 4% of structures required minor edits, respectively. All testing and training data are being made publicly available on The Cancer Imaging Archive.

## Introduction

Artificial intelligence-based auto-segmentation models are being adopted in clinical practice within radiation oncology. The benefits of such approaches are well known and include time-savings^1, 2^, decreases in variability^3^, and quality assurance applications ^4–6^. Such advances are pertinent to the head and neck, as delineation accuracy of OARs (organs at risk) and targets are limited by interobserver and trial protocol variability^7–9^. Within the anatomic site of head and neck, auto-segmentation models developed through deep learning has resulted in a myriad of contouring approaches^4, 10–17^. Many of these models focus on contouring OARs that can be delineated from radiotherapy simulation CT scans. Commercial models for fully automated target segmentation in the head and neck are not yet available, but research in this area is gaining momentum.

Of recent interest is the auto-segmentation of the low-risk, or elective clinical target volume (CTV). The low-risk CTV is comprised of anatomically-defined lymph node-containing regions (“lymph node levels”) that are at risk of metastatic spread, though possess no clinical or radiographic evidence of disease at the time of treatment. The set of lymph node levels selected for inclusion in the low-risk CTV is based on lymphatic drainage patterns from the location of the primary tumor. Generally accepted volumes based on common tumor locations are well documented in consensus guidelines and contouring resources for the head and neck^9, 18^. Automatic CT-based segmentation of these lymph node levels (e.g. I-V) is achievable and has been demonstrated by numerous works^10, 19–25^. Our clinic previously integrated a deep-learning based approach to contour elective lymph node levels in CT scans^10^. This approach groups nodal levels into families (e.g. IA-V, IB-V, II-IV, and retropharyngeal [RP]) so that elective CTVs can be quickly constructed using Boolean algebra for use in manual and automatic treatment planning. However, one significant challenge in the field of target segmentation is keeping pace with changes in clinical practice. Sources of such changes can be driven by changes in contouring guidelines, improvements in image-guidance technology, or evolving evidence that alters our understanding of the balance between toxicity and tumor control^26, 27^. Strijbis et also noted in their work on automated segmentation of levels I-V that contours produced by Cardenas et al are generous, resembling their institution’s PTVs^25^. Based on physician feedback and changes in clinical practice, we sought to develop a new auto-segmentation model that more accurately reflects the narrower treatment volumes utilized in our clinic’s practice today.

Proximal to these CTVs are essential structures which enable deglutition (i.e. swallowing) and mastication (i.e. chewing). When the functions of these structures are compromised, side effects such as dysphagia can occur. Dysphagia is one of the strongest determinants of quality of life following radiation therapy and can affect many physical, mental, and social components of life^28–31^. Many works have well characterized the dose-effect relationships for swallowing and chewing structures^28, 32, 33^. Although publicly available models and repositories exist for OARs in the head and neck, to our knowledge there exists no such repository of swallowing structures. Teguh et al were the last to clinically validate an approach to segment lymph levels and swallowing structures in the head and neck in one combined work, but did so with atlas-based auto-segmentation^19^. Many other authors have studied swallowing and chewing structure segmentation, but only include some of the major swallowing and chewing structures that can be visualized on CT. A comprehensive auto-contouring approach to generate these structures could provide a means for reliable and efficient assessment of dose response studies—especially since manual delineation of these many structures is extremely labor intensive.

The purpose of this work is to present a straightforward approach that can efficiently and effectively create and validate a clinical segmentation tool for 1) individual lymph node levels and 2) swallowing and chewing structures in the head and neck. For the lymph nodes, the hypothesis of this work is that our approach will result in contours that will be as clinically acceptable as physicians’ own manual contours and be preferred at rates equal or superior to manual contours. Such an auto-contouring model could eliminate sources of intra-physician variability present in elective target contouring if used clinically. This will be tested using two double blinded studies to score physician preference and contour quality (for manual vs AI contours). For the swallowing structures, we use a simpler evaluation approach (i.e. no blinded studies) to gauge clinical acceptability of resulting AI contours.

In a busy practice with limited physician time, data scarcity can be a challenge for clinics that are wanting to develop their own tools or perform clinical research. To this end, we have published the first clinically validated dataset of cervical lymph nodes (XXXX) and swallowing and chewing structures (XXXX) on The Cancer Imaging Archive (TCIA).

## Materials and Methods

The MD Anderson Cancer Center has 10 physicians 11 physicists, and 8 dosimetrists who specialize in head and neck cancer at its main hospital. Our treatment planning process for the head and neck uses auto-contouring for organs-at-risk^4^ and lymph node level families^10^ for every patient (> 100 per month). The following subsections outline the data curation, model training, quantitative evaluation, and design of blinded studies. This work followed the recommendations by Baroudi et al which suggest guidance for quantitative (using overlap and distance metrics) and qualitative evaluation (using physician review) of clinical acceptability^34^. Thus, both quantitative and qualitative review metrics are reported from the literature for lymph node segmentation as well as swallowing and chewing structure segmentation.

### Patient data

CTs of 145 patients who were irradiated for a head and neck primary malignancy at MD Anderson Cancer Center were retrospectively curated. All data was gathered under an approved institutional review board protocol. Three CT scanner models were utilized to obtain images: GE Lightspeed/Discovery, Phillips Brilliance Big Bore/64, and Somatom Definition Edge. Modal slice thickness and pixel spacing was 2.5 mm (range: 1.0– 3.3 mm) and 1.17 mm (range: 0.977–1.27 mm), respectively. All patients were imaged supine with head holder and thermoplastic mask specified in our institution’s simulation protocol for head and neck radiotherapy.

The data set of 145 patients were divided into two separate cohorts; one cohort was used to analyze lymph node levels and the other used to analyze swallowing and chewing structures. Forty-seven patients were in the lymph node level cohort. All these patients received definitive radiotherapy without a nodal dissection to minimize anatomic perturbation of the lymph node levels. The remaining 98 patients formed the swallowing/chewing structures cohort.

### Segmentation of ground truth contours

Five radiation oncologists manually contoured seven lymph node levels (IA, IB, II, III, IV, V, RP) on 3 patients each resulting in a total of 105 lymph node levels in 15 patients. These 15 patients served as the testing dataset for the lymph node level segmentation model. Contours were anatomically drawn without margin according to institutional practice.

One radiation oncologist contoured 17 structures involved in swallowing and chewing in the head and neck (tongue, thyroid cartilage, cricoid, cricopharyngeus, glottic area, supraglottic larynx, buccinators, inferior constrictor, medial constrictor, superior constrictor, anterior digastric, posterior digastric, genioglossus, masseter, mylogeniohyoid (mgh) complex, lateral pterygoid, and medial pterygoid) on twenty patients. This cohort was used as the swallowing/chewing structures testing dataset.

### Training and testing methodologies

A three-step methodology was used to create the final multi-class lymph node level segmentation model. First, the publicly available nnUnet model was used to train a multiclass segmentation model using the small, testing dataset of 15 patients^35^. A 3D full resolution model was used with five-fold cross validation; all augmentations were enabled. Left and right contours for each nodal level were combined into one volume to prevent misclassification from left-right flipping augmentations. Second, the model was used to generate lymph node level contours on 32 additional patients (described above) and were edited by a radiation oncology resident. Third, the final model was trained with the 32 patients and tested on the original cohort of 15 patients.

A similar three-step methodology was used to create the swallowing/chewing structures model. First, 20 patients’ structures were manually contoured for use as a standard, and an atlas-based model using Elekta Admire (Elekta AB, Stockholm, Sweden) with batch fusion was created. Second, the atlas was run on the remaining 78 and manually revised by a radiation oncologist. Third, two models were trained to accommodate GPU memory (one with 10 structures and another with 7 structures). The data from the manually revised 78 patients were used for training, and the 20 remaining patients were used for testing. The nnUnet settings to train the models were the same as those mentioned above.

### Quantitative analysis of lymph node level, chewing, and swallowing structures

Dice Similarity Coefficient (DSC) and 95^th^ percentile Hausdorff (HD95th) distance were used as quantitative performance metrics between the ground truth contours and the predicted contours.

For point sets A and B, representing 3-dimensional segmentation volumes, the DSC and HD95^th^ are defined in Equations 1-3. |A| and |B| denote each 3-dimensional segmentation volume; S_1_ and S_2_ are the surfaces from A and B, respectively.

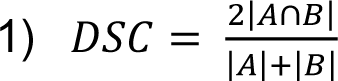

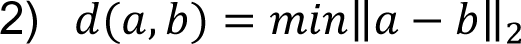

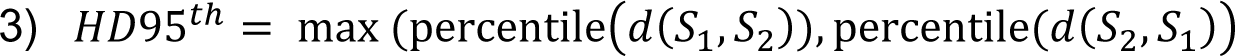

### Blinded studies to evaluate physician preference for lymph node level contours

Two blinded studies were performed to gauge physician preference and quality on the testing dataset. First, ground truth and predicted contour names were identical in the treatment planning system except for a prefix “A” or “B”. This prefix was randomly assigned to the set of human or machine contours to blind the physician to their identity. The colors of the contours in the treatment planning system were also shuffled so that human or machine contours were not the same across all patients. Physicians viewed machine and AI contours simultaneously and scored preference as either A, B, C, or D (A = prefer A, B = prefer B, C = prefer either, or D = prefer neither. Ultimately there were 525 scores collected in the lymph node contour preference dataset, with physicians each evaluating 15 CT scans with 7 lymph node levels contoured (5×15×7). Since a period of over months took place between initial contouring and this blinded study, each of the 5 physicians were unbiased in their scorings, despite having previously contoured ground truth data for 3 of the 15 patients. Thus, physician scores of preference reflect the frequency that their own, the AI, or a colleague’s contours is preferred.

The second blinded study used the same 15 patient cohort. The goal of this blinded study was to quantify the degree to which contours require editing before clinical use. Quantitative scores and comments for each contour (AI and manual) were collected for these patients. The scoring of the manual contours provided a control and comparison for the AI contours. Due to the labor-intensive nature of this task, each physician only rated 3 patients each. One of the three patients was originally contoured by the scoring physician; two of the three patients were originally contoured by a different physician. A 5-point Likert scale was used:

1. Unusable: The automatically generated contours are unusable (ie, wrong body area, outside confines of body, etc).
2. Major edits are necessary: Edits that the reviewer judges are required to ensure appropriate treatment are present. Edits required are significantly substantial and the user would prefer to start from scratch.
3. Minor edits are necessary: Edits that the reviewer judges are clinically important exist. Also, it is more efficient to edit the automatically generated contours than start from scratch.
4. Minor edits are not necessary: Stylistic differences exist, but differences are not clinically important. The current contours are acceptable.
5. Use-as-is: Clinically acceptable, could be used for treatment without change.

Student’s t-tests were used to quantify whether the mean of the clinical acceptability scores were significantly different for manually generated versus AI generated contours.

### Qualitative scoring for swallowing and chewing structures

The aforementioned five-point Likert scale was used to qualitatively score structure predictions on the 20-patient test set from the swallowing/chewing structures patient cohort. Clinically acceptability was scored by one physician.

## Results

### Lymph node auto-segmentation performance

Median DSC for the test set (n = 15) were 0.83, 0.89, 0.88, 0.85, 0.83, 0.79, 0.77 for levels IA, IB, II, III, IV, V, and RP, respectively. Median HD95^th^ (in mm) are 2.5, 2.7, 3.3, 4.2, 5.3, 5.5, and 2.9 for levels IA, IB, II, III, IV, V, and RP, respectively. DSC’s are featured in Table I for comparison to other recent lymph node segmentation approaches in the literature.

### Physician preference for AI-based lymph node segmentations

Across all contours, physicians preferred the AI contours at a rate of 247/525 = 47%. The manual contours were preferred at a rate of 88/525 = 16.8%. The AI contour was superior to or equally preferred to the manual contour at a rate of 436/525 = 83%. This rate is the sum of “AI” and “Either” (Table 1). For levels IA, IB, II, III, IV, V, and RP, the AI contour was superior to or equally preferred to the manual contour at rates ranging from 75% to 91%. One physician scored only 1 of 525 contours (level IV) as preferring neither the AI nor the MD contour. In this instance, the physician that made the ground truth manual contour and the physician that scored were different physicians. On average, physicians preferred their original contours (when unknowingly reviewing their own contours which were blinded) over that of the AI contours only 18% of the time (range = 0 - 29%).

**Table 1.** | Physician preference scoring for manual and AI contours

|  | IA | IB | II | III | IV | V | RP | Sum |
| --- | --- | --- | --- | --- | --- | --- | --- | --- |
| Either | 60% | 45% | 29% | 44% | 28% | 19% | 27% | 36% |
| AI | 24% | 40% | 51% | 44% | 47% | 60% | 64% | 47% |
| Manual | 16% | 15% | 20% | 12% | 24% | 21% | 9% | 17% |
| Neither | 0% | 0% | 0% | 0% | 1% | 0% | 0% | 0% |
| AI or Either | 84% | 85% | 80% | 88% | 75% | 79% | 91% | 83% |

**Table 2.** | Physician scoring of clinical acceptability for manual and AI contours

|  | IA | IB | II | III | IV | V | RP | All |
| --- | --- | --- | --- | --- | --- | --- | --- | --- |
| AI | 4.2 (3-5) | 4.4 (4-5) | 4.2 (3-5) | 4.4 (4-5) | 4.1 (3-5) | 4.1 (3-5) | 4.5 (3-5) | 4.3 (3-5) |
| Manual | 4.4 (3-5) | 4 (3-5) | 3.9 (3-5) | 4.1 (3-5) | 3.9 (3-5) | 3.9 (2-5) | 3.7 (3-4) | 3.9 (2-5) |
| p-value | 0.37 | 0.07 | 0.06 | 0.11 | 0.42 | 0.42 | <0.01 | <0.01 |

### Swallowing and chewing structures auto-segmentation performance

### Physician review of AI-based swallowing and chewing structure segmentations

Of the 340 contours reviewed, 100% were found to be 3 or greater (minor edits - use as is) as noted in Table 3. 96% percent of contours were scored as 4 or 5 (stylistic differences, use-as-is). Thyroid cartilage, glottic area, larynx, buccinators, digastrics, masseters, mgh complex, and pterygoids had perfect scores (5’s) across all patients. Four common issues were identified (1) The genioglossus would contour into the mgh complex when transitioning, (2) The anterior digastrics overcontour into the mgh complex, (3) The anterior digastrics would overcontour posteriorly, and (4) Transitions between the superior constrictor and medial constrictor, medial constrictor and inferior constrictor, and inferior constrictor and cricopharyngeus would be incomplete by missing a slice between structures or by splitting into islands at the transition. In general, the algorithm performed well in the presence of dental artifact (Fig A1). Examples of minor edits needing to be performed are featured in Figure 6 in the presence of tumor or dental stent which distorts the tongue.

**Figure 1.**
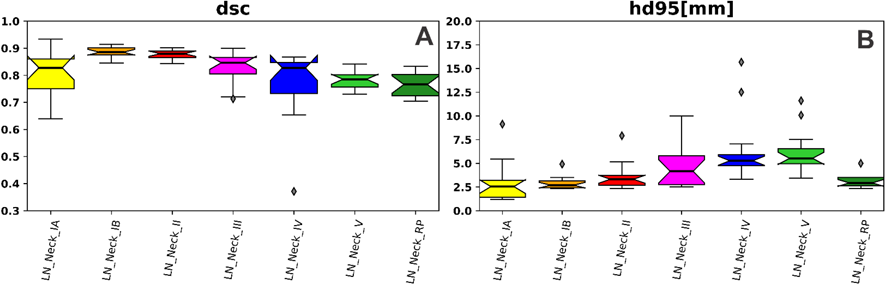
| Quantitative metrics for lymph node level segmentation. dsc, dice similarity coefficient; hd95, 95^th^ percentile Hausdorff distance

**Table 3.**
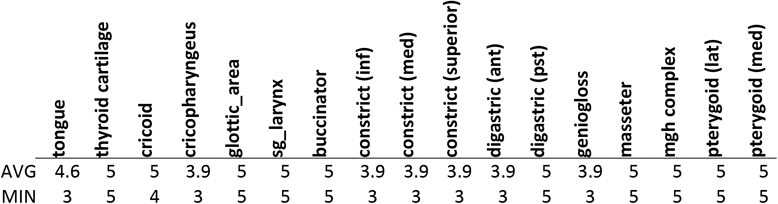
| Physician scoring of clinical acceptability for swallowing and chewing structures

|  | tongue | thyroid cartilage | cricoid | cricopharyngeus | glottic_area | sg_larynx | buccinator | constrict (inf) | constrict (med) | constrict (superior) | digastric (ant) | digastric (pst) | geniogloss | masseter | mgh complex | pterygoid (lat) | pterygoid (med) |
| --- | --- | --- | --- | --- | --- | --- | --- | --- | --- | --- | --- | --- | --- | --- | --- | --- | --- |
| AVG | 4.6 | 5 | 5 | 3.9 | 5 | 5 | 5 | 3.9 | 3.9 | 3.9 | 3.9 | 5 | 3.9 | 5 | 5 | 5 | 5 |
| MIN | 3 | 5 | 4 | 3 | 5 | 5 | 5 | 3 | 3 | 3 | 3 | 5 | 3 | 5 | 5 | 5 | 5 |

## Discussion

This work has demonstrated how a publicly available deep learning model trained with expertly curated data (n = 32 lymph node dataset, n = 78 swallowing/chewing structure dataset) can result in a practical segmentation tool that is both accurate and clinically acceptable. Furthermore, this work has resulted in two published datasets (in The Cancer Imaging Archive) that are available for clinical researchers to reproduce or build upon the field of head and neck CT-based tissue segmentation. To our knowledge, these two datasets represent the first publicly available datasets for swallowing and chewing structures and lymph nodes contoured on non-contrast CT, both with physician scores of clinical acceptability.

One early work in lymph node and swallowing structures segmentation in the head and neck was by Teguh et al which obtained a DSC of 0.73 on average for lymph node levels I-V and 0.50–0.71 DSC for swallowing and chewing structures^19^. Since then, multiple authors have continued to build more accurate models using deep learning, while simultaneously increasing the number of structures predicted by deep learning models (Tables 4 and 5). While various works such as van der Veen and Weissmann et al reported qualitative evaluations (e.g. radiation oncologist scores) and quantitative evaluations (e.g. DSC, HD) for lymph node model contours, few authors reported what proportion of deep learning contours require edits before the contours can be used clinically (Table 4). For swallowing and chewing structures, most authors have reported what percentage of their test set contours require edits before clinical use (Table 5). These rates have ranged from 100% to 18% for various structures with DSC’s ranging from 0.5 to at most 0.91 on average or median. In addition to accuracy and clinical acceptability, previous works have extensively studied human interobserver variability in lymph node level contouring and have quantified it in terms of DSC^23, 24^. Weissman et al demonstrated that when physicians re-contoured their own volumes, DSC was 0.77 on average^24^. Overall, they reported that the accuracy of their deep learning approach performed with 0.78 DSC on average across 20 nodal levels and made their deep learning model publicly available.

**Table 4.** | Lymph node level segmentation performance in the literature

| Authors | Year | Approach | volume | minor edits or greater required | DSC |
| --- | --- | --- | --- | --- | --- |
| Teguh et al<sup>19</sup> | 2011 | multi-atlas | I-V | NA | 0.73 |
| Yang et al<sup>21</sup> | 2014 | multi-atlas | low-risk CTV | NA | 0.78 |
| Wong et al<sup>22</sup> | 2020 | deep learning | IB, II, III, IV, V, RP (as one volume) | NA | 0.72 |
| Cardenas et al<sup>10</sup> | 2020 | deep learning | Ia-V, Ib-V, II-IV, and RP | 43% | (0.81 - 0.90) |
| van der Veen<sup>23</sup> | 2020 | deep learning | 17 nodal levels | 100%* | (0.46 - 0.82) |
| Strijbis et al<sup>25</sup> | 2022 | deep learning | I, II, III, IV, V | NA | (0.71 - 0.85) |
| Weissmann et al<sup>24</sup> | 2022 | post processing | 20 nodal levels | NA^ | 0.78 |
| **Maroongroge et al** | **2023** | **deep learning** | **IA, IB, II, III, IV, V, RP** | **8%** | **(0.77 - 0.89)** |

\* edits were 1.4 mm on average
^ Contours were scored by a clinical expert on a 0-100 scale, and was 81 on average.

**Table 5.** Swallowing and chewing structures segmentation performance in the literature

| Authors | Year | Approach | volume | minor edits or greater required | DSC |
| --- | --- | --- | --- | --- | --- |
| Teguh et al<sup>1</sup> | 2011 | multi-atlas | swallowing/chewing structures | NA | (0.50 - 0.71)<sup>^^</sup> |
| van der Veen et al<sup>2</sup> | 2019 | deep learning | HN OARs and constrictor muscles | NA<sup>^</sup> | (0.71 - 0.97) |
| van Dijk et al<sup>3</sup> | 2020 | deep learning | 8 Upper digestive and airway-related* | ~18-59% | (0.52-0.91) |
| Li et al<sup>4</sup> | 2022 | deep learning | 4 swallowing structures | < 50% | (0.60-0.84) |
| Iyer et al<sup>5</sup> | 2022 | deep learning | 4 swallowing/chewing structures* | (23-100%) | (0.69–0.87) |
| **Maroongroge et al** | **2023** | **deep learning** | **17 swallowing/chewing structures\*** | **4%** | **(0.86-0.96)** |

<sup>^</sup> 100%, < 2mm MSD on average
\*left/right designation not included in count
<sup>^^</sup> atlas contours vs reference contours

Regarding the lymph nodes segmentation performance of our approach, quantitative analysis of the lymph node level segmentations indicated that DSC ranged from 0.77 to 0.89 across all levels and is comparable to other recent approaches in the literature (Table 4). Regarding qualitative evaluation, a blinded study investigating physician preference indicated that the AI contours were superior to or equally preferred to the manual contours at rates ranging from 75% to 91%. In addition, a blinded study of clinical acceptability indicated that there was no significant difference in clinical acceptability between manual contours and AI contours for levels IA, IB, II, III, IV, and V, with greater clinical acceptability demonstrated for AI-generated RP contours. Furthermore, only 8% of AI lymph node contours require edits for clinical use as determined by our team of sub-specialized head and neck radiation oncologists. Thus, the hypothesis of this work can be accepted, since 1) there is no difference in clinical acceptability between manual and AI contours and 2) the AI contours were superior to or equally preferred to the manual contours. Although not directly comparable, our average DSC’s in the nodal test set are all greater than previously reported measures of interobserver variability mentioned above. Anecdotally, this may suggest that our model accuracy has similar or smaller levels of variance compared to human contouring in our practice since there was no preference for manual vs AI contours (p <0.01) for all but one nodal level. As is performed in clinical practice, the lymph node model learned to seamlessly abut adjacent nodal levels (i.e. no gaps). However, this abutment did not coincide with the axial slice planes which are orthogonal to the superior-inferior direction (Figure 2B). A slice plane adjustment function was developed by Weissman et al, to post process contours so that they abut in axial planes and more closely resemble human contours which are typically contoured in axial views^24^. While this technique resulted in better physician ratings in Weissman’s study (vs no post processing), such a technique was not applied to our testing cohort, as our lymph node contours were scored as equivalent or superior to manual contours without post processing.

**Figure 2.**
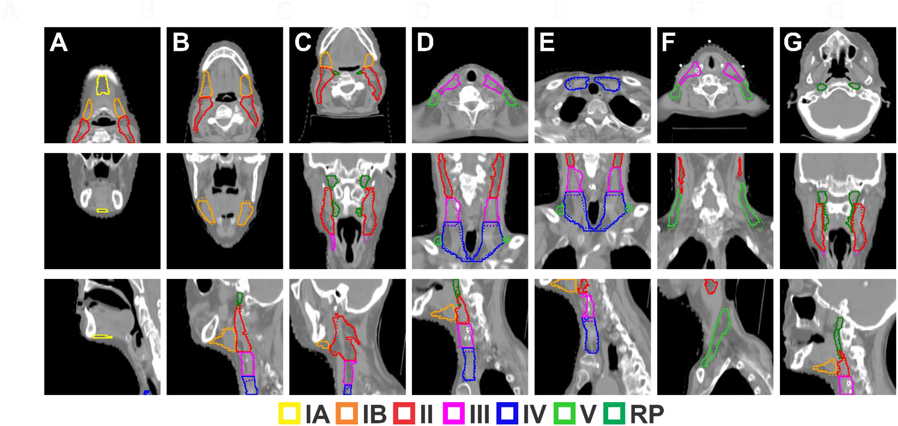
| Segmentations of lymph node levels IA, IB, II, III, IV, V, and RP (columns A-G, respectively). Solid lines represent ground truth segmentations and dotted lines represent predicted segmentations.

Strijbis et al demonstrated similar quantitative lymph node model performances values to our work but used a training cohort roughly twice the size of our training cohort^25^. However, as demonstrated by Yu et al, there are diminishing returns for increasing U-Net segmentation performance as a function of training dataset size^36^. This is especially true when the training data (in this case CT) contain images of patients with consistent patient positioning, immobilization, and imaging protocol. Weismann et al demonstrated that a clinically acceptable lymph segmentation model could be made with the publicly available nnUnet architecture with a small cohort of patients (n = 35)^24^. Likewise, we have found this number to be adequate for our patient population of head and neck patients which receive radiotherapy simulation CTs.

Regarding the swallowing structures segmenation performance, excellent quantitative and qualitative measures were obtained on the swallowing/chewing structures test set. Our work produced a model that segments 17 different swallowing and chewing structures, more than any work, to our knowledge, has produced. This publicly available data will be a valuable source of data for future head and neck studies. Median DSC was greater than 0.86 for all structures, and although values in the literature cannot be directly compared between different datasets, our approach yields the most accurate structures reported in terms of DSC. Furthermore, it was demonstrated that 96% percent of contours in the test set could be used clinically without edit and this percentage is much greater than any rates previously reported in the literature for swallowing and chewing structures (Table 5). Dental stents distorting tongue position (Figure 6B) and the presence of abutting tumor (Figure 6B) were common factors which necessitated minor edits. The hypothesis of this work, that swallowing and chewing AI contours are clinically acceptable, can be accepted.

One limitation of this work is that the impact on the model that variations in patient population and radiotherapy simulation protocol were not studied. Future studies would benefit from studying the effects of patient positioning, adenopathy (as noted in Fig 3A), nodal level dissection, bulky disease, and presence of contrast upon segmentation performance. In this work, training and testing cohorts for both datasets used a three-step methodology in which ground truth data was generated by a sparsely trained deep learning model or atlas-based model. Although this strategy is useful for perpetuating sparse data, it can bias the curation process if contours are not comprehensively edited after it is generated by the atlas or sparsely trained model. Lastly, not all nodal levels were included in this work, as many listed in consensus guidelines are not commonly used. Future model refinements will include a more comprehensive selection of nodal levels.

**Figure 3.**
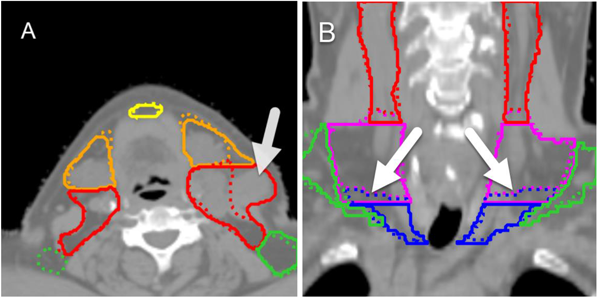
| Featured segmentations. White arrow in A indicates bulky adenopathy at level II (red). White arrows in B indicate where predicted levels III (dotted blue) and IV (dotted pink) slope in the inferior-superior direction.

**Figure 4.**
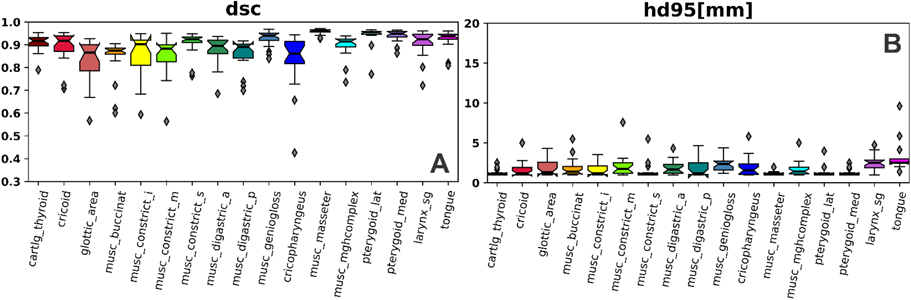
| Quantitative metrics for swallowing/chewing structure segmentations. dsc, dice similarity coefficient; hd95, 95^th^ percentile Hausdorff distance Median DSC for the test set (n = 20) ranged from 0.86 to 0.96 (Fig 4A); median HD95^th^ (in mm) ranged from 1.2 to 2.5 (Fig 4B). Median DSC was greater than 0.90 for 11/17 structures.

**Figure 5.**
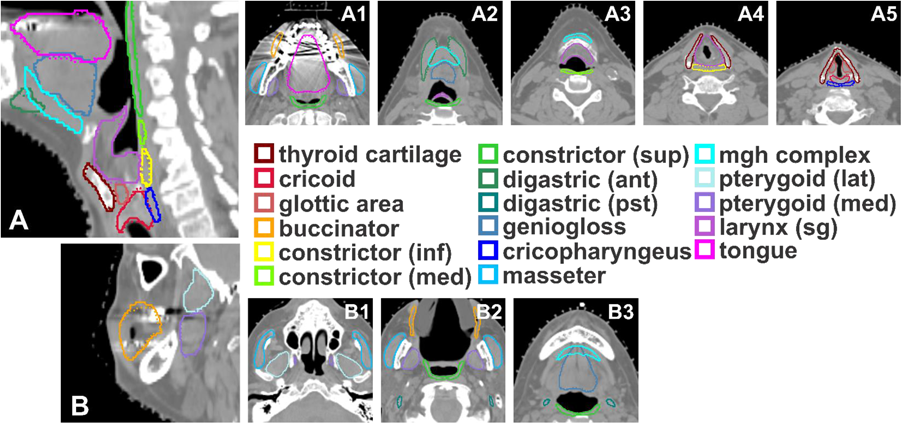
| Segmentations of all swallowing/chewing structures. 5A is the mid-sagittal plane that cuts through a majority of the midline structures; 5B is a sagittal plane that cuts through the buccinator and pterygoids. Solid lines represent ground truth segmentations and dotted lines represent predicted segmentations. In many of the figures, solid and dotted lines are overlapped.

**Figure 6.**
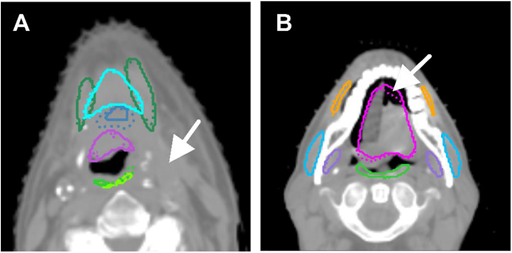
| Featured segmentations for swallowing/chewing structures requiring minor edits. White arrow in A indicates the presence of tumor that is displacing local anatomy. White arrows in B indicate where the tongue is displaced by the dental stent.

These models for lymph nodes and swallowing and chewing structures are being integrated into our clinical practice at MD Anderson as well as the Radiation Planning Assistant (RPA), a web-based, FDA 510k cleared platform that provides contouring and planning to clinics with low resources. Future work will involve multi-institutional studies to evaluate robustness as well as application based-works that use these tools for clinical trial quality assurance and toxicity analyses.

## Conclusion

This work demonstrated an approach to generate clinically acceptable contours for lymph node levels, swallowing structures, and chewing structures for CT-based segmentation in the head and neck. A publicly available deep learning model using data curated by sub-specialized radiation oncologists in the head and neck resulted AI contours which are, on the vast majority, clinically acceptable and highly accurate. This work has also produced two separate datasets that are the first publicly available datasets for lymph node levels and swallowing and chewing structures in the head and neck.

## Data Availability

All data are being made publicly available through repositories.

